# Disclosure and Double Standards: A Mixed Methods Study of Self-Disclosure of Mental Illness or Addiction Among Medical Learners

**DOI:** 10.1101/2023.05.16.23290047

**Authors:** Aliya Kassam, Benedicta Antepim, Javeed Sukhera

**Author notes:** Corresponding Author: Dr. Aliya Kassam, PhD Research Lead, Office of Postgraduate Medical Education Associate Professor, Department of Community Health Sciences Cumming School of Medicine 3330 Hospital Drive NW Calgary, AB, Canada T2N 4N1 T F E.

## Abstract

**Purpose:** Despite the proliferation of initiatives to address wellbeing and reduce burnout, mental illness and addiction stigma remains rooted within medical education and healthcare. One mechanism to address this stigma is self-disclosure. Given the paucity of literature on self-disclosure in medical learners, we sought to explore perceptions of self-disclosure in medical education.

**Method:** In a mixed method, convergent triangulation design, authors recruited medical learners from across Canada. Quantitative data included the Opening Minds Scale for Healthcare providers (OMS-HC), the Self Stigma of Mental Illness Scale (SSMIS), a wellbeing measure, and questions regarding substance use from Statistics Canada. Qualitative data included semi-structured interviews, which were collected and analyzed using a phenomenological approach. Data were collected simultaneously, analyzed separately, and then triangulated. Discrepancies were discussed until consensus was achieved.

**Results:** Overall, N= 125 medical learners (n= 67 medical students, n=58 resident physicians) responded to our survey, and N=13 participated in interviews (n = 10 medical students, n =3 resident physicians). OMS-HC scores showed resident physicians had more negative attitudes towards mental illness and disclosure (47.7 vs. 44.3, *P* = 0.02). Self-disclosure was modulated by the degree of intersectional vulnerability of the learner’s identity. When looking at self-disclosure, people who identified as men had more negative attitudes than people who identified as women (17.8 vs 16.1, *P* = 0.01). Racialized learners scored higher on self-stigma. Interview data suggested that disclosure was fraught with tensions, but perceived as having a positive outcome, including the perception that self-disclosure made learners better physicians and educators in the future.

**Conclusion:** The individual process of disclosure is complex and appeared to become more challenging over time due to the internalization of negative attitudes about mental illness. Intersectional vulnerability in medical learners warrants further consideration. Fear of disclosure is an important factor shaped by the learning environment.

---

Medical education can take place in a highly stressful environment, particularly for individuals with lived experience of mental illness or addiction. Despite the high rates of mental illness or addiction in medical learners, many individuals do not seek help for fear that the stigma of these conditions will tarnish their reputation and careers.^1-3^ The consequences of unaddressed mental illness or addiction among medical learners are potentially disastrous. For example, a recent Canadian survey found medical students had significantly higher rates of psychological distress, suicidal ideation, and mood and anxiety disorders compared to the general population.^1^ Similar data mirrors these trends in the United Kingdom, Europe, and jurisdictions outside North America.^2^ Any efforts to enhance the wellbeing of medical learners will remain limited unless the role of stigma in the clinical learning environment is addressed in a meaningful way.

Stigma includes stereotypes (negative beliefs), prejudice, and discrimination.^3^ Stigma can be experienced towards others, or towards oneself.^4,5^ Self-stigma can have negative consequences including a loss of self-efficacy, and when internalised, self-stigma can also contribute to a sense of learned helplessness.^4,5^ Previous research on stigma suggests that stigma is enacted through dynamic processes that include individual, organizational, and societal level influences. ^3,5^ In a recent study of stigma amongst medical learners, stigma against help seeking was counterbalanced by the ideal of heroic disclosure which was perceived as challenging within a system that upholds cultural norms of perfectionism and invulnerability.^6^

Interventions to reduce self-stigma therefore often promote self-disclosure.^4^ The concept of self-disclosure is defined as a process through which individuals verbally reveal themselves to others.^7^ By sharing one’s experience, self-disclosure helps to bring stigmatized conditions such as mental illness or addictions out of the shadows, disconfirms stereotypes, and normalizes help-seeking. Therefore, self-disclosure may have potential benefit in improving the wellbeing of medical learners. ^3,5^

Current knowledge on self-disclosure in medical education is limited. Existing literature includes a few studies that highlight the value of faculty self-disclosure^8^; and evidence that those who disclose are less likely to receive an interview invitation.^9^ There have been studies that explore attitudes on the topic^10,11^ however, there have been little no empirical investigations of the process of learner self-disclosure including information about how disclosure relates to self-stigma, learners’ social identity, and the barriers or enablers for those who end up disclosing. A deeper understanding of self-disclosure among medical learners may help advance our understanding of both how stigma manifests in the medical learning environment and how to address it.

The aim of our study was to explore perceptions of self-disclosure in medical education. We sought to explore barriers and enablers of self-disclosure related to knowledge, attitudes, and behaviour^3^, through a deeper understanding of the perceived consequences (both positive and negative) of self-disclosing among medical learners. Our overarching research question was, what is the experience of medical learners regarding self-disclosure of mental illness or addictions? We also sought to understand the following sub-questions: (1) What factors enable or constrain self-disclosure behaviour in medical learners? (2) What are the perceived positive and negative consequences of self-disclosure in medical learners? (3) How do medical learners perceive experiences of self-disclosure?

## Method

We used pragmatism as the theoretical construct for our research study. Pragmatism assumes that knowledge is created from socially shared experiences and is influenced by our beliefs, habits, and interactions with others.^12^ In our study, we first identified a common problem among medical learners pertaining to the stigma of mental illness or addiction and perceptions of self-disclosure.

### Design

This study used mixed methods which is commonly used in pragmatic research, by collecting and analyzing data with a convergent triangulation research design.^13^ Our rationale for using a mixed methods design was to explore the construct of self-disclosure through complementary yet distinct sources of data. A mixed methods design was useful for our study because little is known regarding self-disclosure in medical learners, and therefore complementary methods will help inform a conceptual framework that can become the basis for future work in this area. By using a convergent triangulation design, our study included methodological borrowing of distinct, yet complementary epistemologies associated with hermeneutic and post-positivist orientations. This mixing of methodologies is intentional and aligns with the concept of methodological borrowing as described by Varpio and colleagues.^14^ This research was approved by the Conjoint Health Research Ethics Board at the University of Calgary (ID:20-1149) and the Office of Human Research Ethics (OHRE) at Western University (ID: 116634).

### Setting and participants

Canada is home to 17 diverse medical schools spanning a wide geography. Each school varies regarding curricular approach and format. The study team had support and infrastructure at two sites in Canada (each in western Canada and eastern Canada) where ethics review was sought. Data collection took place across the 17 medical schools. To ensure the highest level of confidentiality and protection of sensitive information we had an interviewer who was arms-length (author B.A.) who had no influence on the participants’ training or career progression.

### Data Collection

Data was collected between August 2020 and August 2021 and analyzed in 2022. Recruitment for the study took place at the respective co-principal investigators home institutions through internal communication channels such as listservs. All other medical schools in Canada were recruited by snowball sampling through email as well as Twitter.

#### Quantitative Arm (QUAN)

The inclusion criteria for participation in the quantitative survey was any current medical learner (medical student or resident) who had experience of a mental illness or addiction.

Quantitative data was collected using the Qualtrics online survey tool.^15^ Our survey consisted of several previously developed and psychometrically tested questionnaires.^16-18^ Supplemental material shows the validated questionnaires used. ^16-18^ We also included questions pertaining to substance use over the past one year from Statistics Canada. All continuous quantitative data were tested for assumptions of normality and where data was positively skewed, data was log transformed and geometric means were computed. Continuous data were analyzed using independent samples t-tests comparing mean scores and thus data for demographic variables was dichotomized. Effect sizes in the form of Cohen’s *d* were also computed to determine how meaningful the relationship between groups were.

#### Qualitative Arm (QUAL)

The inclusion criteria for the participation in the qualitative interview was any current medical learner (medical student or resident) who had self-disclosed their mental illness or addiction in training. Interview participants were recruited from survey participants by asking them to contact complete a form with their contact information should they choose to participate in the interview. In seeking a rich understanding of the factors that influence self-disclosure of mental illness or addiction for medical learners, we chose to inform the qualitative arm of our study with phenomenological inquiry as a methodology. Phenomenology, particularly hermeneutic phenomenology explores the meaning of lived experience and the contextual forces that shape an individual’s experiences.^19^ Phenomenological approaches have been utilized for similar explorations in related topics in medical education such as shame experiences^20^, and microaggressions^21^ in clinical learning environments. A hermeneutic approach directs attention to an experience as it is lived, rather than how it is conceptualized. Hermeneutic phenomenology is suited to our research topic and emphasizes the limitations of “bracketing” for the research team.^19^ Our method of analysis of the qualitative data was informed by phenomenological approaches to analysis^19^. Such approaches allow researchers to reveal the deeper meanings of the experiences being studied and to identify the unique experiences of the participants perceptions of self disclosure of mental illness or addiction in medical education. During our analysis we openly reflected on, shared, and attended to subjectivity during an iterative process of data collection, reflection, reflexivity, and analysis.^22^

We conducted qualitative analysis on both open comments in the survey and throughout the semi-structured interviews. Two investigators (J.S. and B.A.) developed a semi-structured discussion guide based on existing literature on self-disclosure. Questions focused on eliciting narratives of experience. We asked about each participant’s experience of disclosure including predisposing factors, motivations to disclosure, reactions from self, reactions from others, and perceived meaning of disclosure for their future career as a physician and/or educator. We scheduled interviews for 90 minutes to allow for deeper probing and reflection on participant experiences consistent with a phenomenological approach. Interviews were audio-recorded and transcribed verbatim as well as de-identified before analysis.

Data were analyzed in accordance with the tenets of hermeneutic phenomenological analysis described in the literature.^19,23^ This approach identifies participants’ interpretations and first-order constructs which are subsequently integrated with the researcher’s second order constructs. Stages include immersion, understanding, abstraction, synthesis, illumination, and integration.^19^ Both J.S. and B.A. consensus coded the first several transcripts and then J.S. conducted independent coding which was discussed at regular intervals with other members of the team.

### Reflexivity

Our team acknowledged that methodological borrowing requires that scholars be competent in both methodologies.^14^ Both co-primary investigators (A.K. and J.S.) have doctorates in health services research and medical education respectively, are full-time academics who are well-versed in both qualitative and quantitative research methods as well as share an interest in mental health or addiction. Author J.S. is also trained as a psychiatrist and A.K. is trained as a psychiatric epidemiologist/health services researcher. They work separately in distinct academic medical centers that teach medical learners.

In conducting this research, both investigators and research team members intended to consistently and iterative test their own assumptions and the framing of the issue of self-disclosure of mental illness or addiction based on their own personal and professional experiences. Since the investigators have previous knowledge of mental illness, they will be aware that if there are findings from a participant that are unexpected or do not fit with preconceived assumptions of ideas, that this is an opportunity to reflect on these previous assumptions. When analyzing interviews, we sought to remain open and non-judgmental. Given that the data is largely a result of the interaction between the investigators and the participant in interviews, investigators and research sought ongoing critical awareness of this relation.

## Results

### Quantitative (QUAN) – Survey

Overall, N= 125 medical learners (n= 67 medical students, n=58 resident physicians) responded to our survey. Table 1 shows the demographic data of the survey participants. Of the n=77 participants that responded to the survey question about disclosure, 36.3% reported having disclosed a mental illness or addiction within their learning environment whereas 24.7% have only considered disclosing a mental illness or addiction. Due to non-applicable responses and missing data, not all percentage estimates will total 100.

**Table 1.**
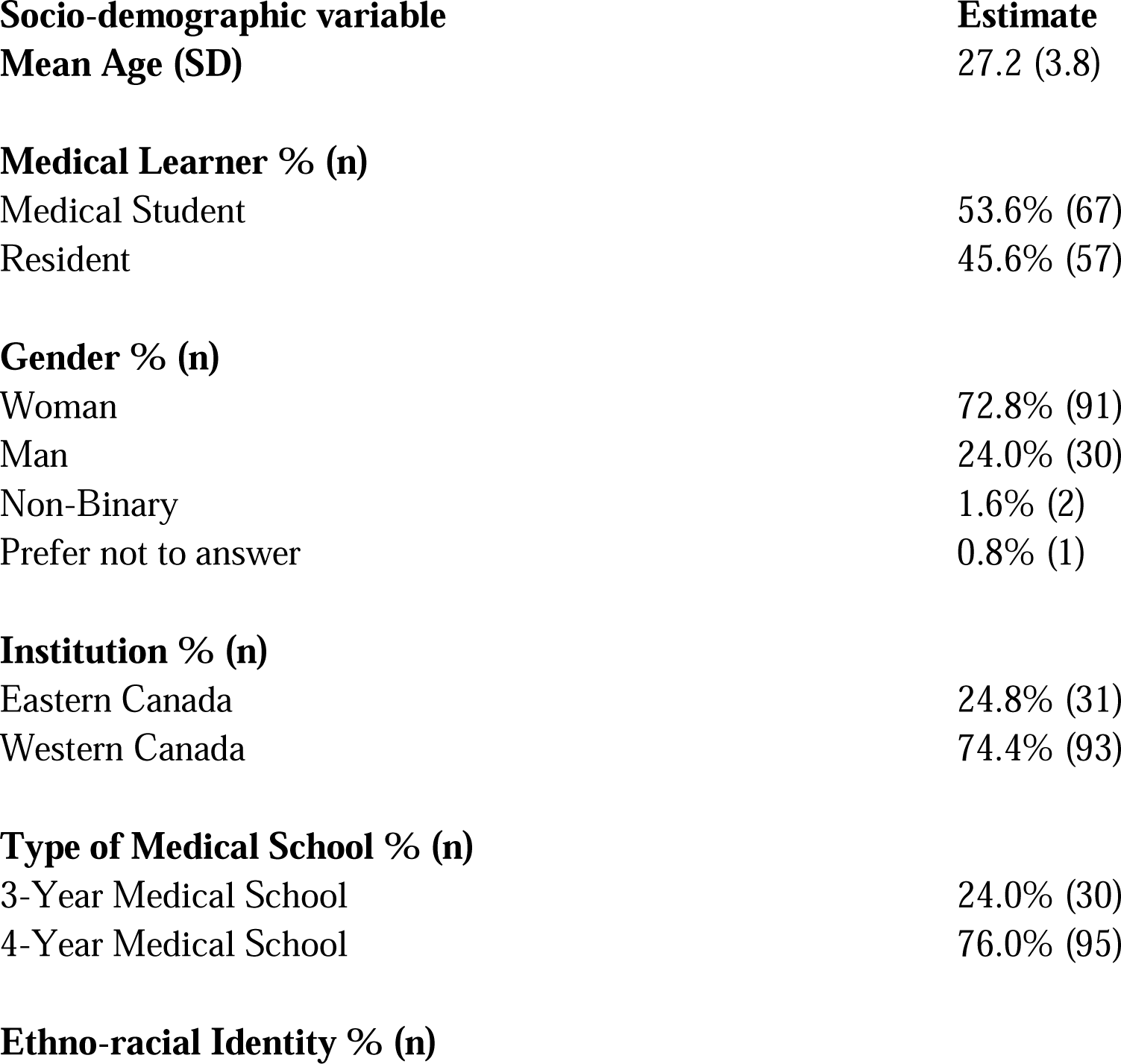
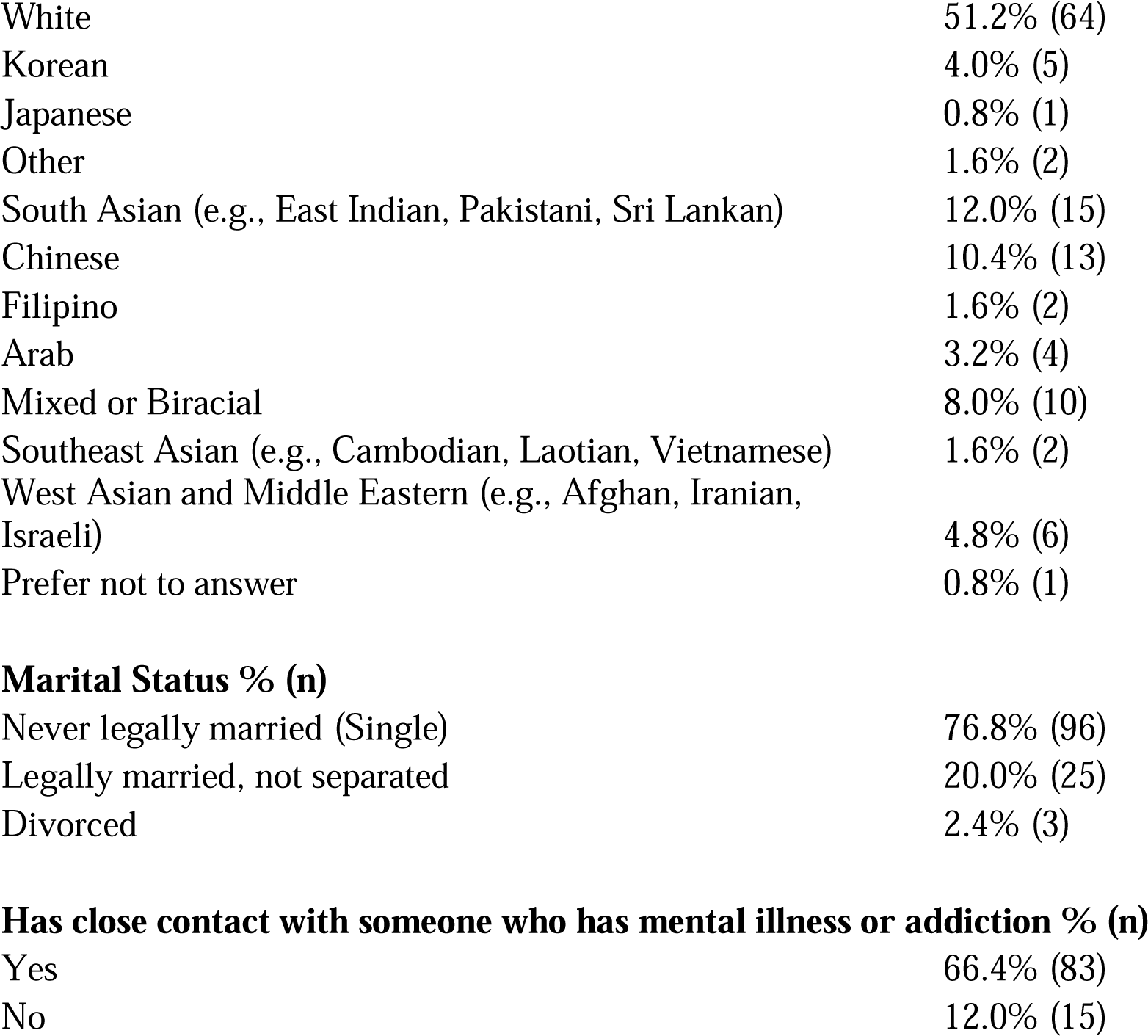
Demographic Characteristics of Survey Participants.

| **Socio-demographic variable** | **Estimate** |
| --- | --- |
| **Mean Age (SD)** | 27.2 (3.8) |
| **Medical Learner % (n)** |  |
| Medical Student | 53.6% (67) |
| Resident | 45.6% (57) |
| **Gender % (n)** |  |
| Woman | 72.8% (91) |
| Man | 24.0% (30) |
| Non-Binary | 1.6% (2) |
| Prefer not to answer | 0.8% (1) |
| **Institution % (n)** |  |
| Eastern Canada | 24.8% (31) |
| Western Canada | 74.4% (93) |
| **Type of Medical School % (n)** |  |
| 3-Year Medical School | 24.0% (30) |
| 4-Year Medical School | 76.0% (95) |
| **Ethno-racial Identity % (n)** |  |
| White | 51.2% (64) |
| Korean | 4.0% (5) |
| Japanese | 0.8% (1) |
| Other | 1.6% (2) |
| South Asian (e.g., East Indian, Pakistani, Sri Lankan) | 12.0% (15) |
| Chinese | 10.4% (13) |
| Filipino | 1.6% (2) |
| Arab | 3.2% (4) |
| Mixed or Biracial | 8.0% (10) |
| Southeast Asian (e.g., Cambodian, Laotian, Vietnamese) | 1.6% (2) |
| West Asian and Middle Eastern (e.g., Afghan, Iranian, Israeli) | 4.8% (6) |
| Prefer not to answer | 0.8% (1) |
| **Marital Status % (n)** |  |
| Never legally married (Single) | 76.8% (96) |
| Legally married, not separated | 20.0% (25) |
| Divorced | 2.4% (3) |
| **Has close contact with someone who has mental illness or addiction % (n)** |  |
| Yes | 66.4% (83) |
| No | 12.0% (15) |

All continuous variables were positively skewed except for the WHO-5 wellbeing score, Attitudes towards Disclosure subscale score and the total score of the Opening Minds Scale for Healthcare Providers (OMS-HC) shown in Table 2. Medium effect sizes (0.3-0.5) were found with racialized learners having more stigmatizing attitudes overall as well as with respect to disclosure. Medium effect sizes were also found with learners identifying as women having less stigmatizing attitudes overall and with respect to disclosure. Large effect sizes (0.8 and above) were found with learners who had close contact with a person with mental illness or addiction having less stigmatizing attitudes overall as well as with respect to disclosure. Overall scores of the WHO-5 showed poor wellbeing across the participants with a mean total score of 12.5 (95% CI 11.5-13.5). Medical learners who were married and not separated had statistically significantly higher wellbeing scores than those who were single (14.3 vs. 12.2, *P*=0.03). Across all other stratified groups of medical learners however, wellbeing scores prompted a signal to screen for depression given that participants scored 13 or below.

On the OMS-HC, resident physicians had more negative attitudes towards people with mental illness and disclosing a mental illness (47.7 vs. 44.3, *P* =0.02). When looking at the Attitudes towards Disclosure subscale of the OMS-HC, people who identified as men had more negative attitudes than people who identified as women (17.8 vs 16.1, *P* = .01).

**Table 2.** Non-skewed Measures of Wellbeing, Attitudes Towards Mental Illness (MI) and Attitudes Towards Disclosure of MI.

|  | Measure |  |  |
| --- | --- | --- | --- |
|  | WHO-5 | OMS-HC – Total Score | OMS-HC- Disclosure /Help-Seeking Subscale |
| Demographic Variable | Mean (SD, 95% CI) | Mean (SD, 95% CI) | Mean (SD, 95% CI) |
| Third Year Medical School | 12.6 (5.0, 11.5-13.8) | 46.1 (7.3, 44.4-47.8) | 16.5 (3.3, 15.7-17.3) |
| Four Year Medical School | 12.3 (4.9, 10.0-14.5) | 45.4 (6.0, 42.4 – 48.4) | 16.4 (3.3, 14.8-17.9) |
| Cohen's *d* Effect Size | 0.08 | 0.09 | 0.05 |
| Eastern Canada | 12.7 (5.1, 11.5-13.8) | 44.9 (5.9, 42.1-47.7) | 16.1 (3.2, 14.7-17.6) |
| Western Canada | 12.5 (4.9, 10.0-14.5) | 46.2 (7.3, 44.5-48.0) | 16.6 (3.3, 15.8-17.4) |
| Cohen's *d* Effect Size | 0.03 | 0.19 | 0.14 |
| Medical Student | 12.7 (5.5, 11.2- 14.3) | **44.3 (6.6, 42.3-46.2)<sup>b</sup>** | 16.1(3.1, 15.2-17.0) |
| Resident | 12.4 (4.3, 11.1-13.6) | **47.7 (7.1, 45.6-50.0)<sup>b</sup>** | 16.9 (3.5, 15.8-17.9) |
| Cohen's *d* Effect Size | 0.08 | 0.50 | 0.22 |
| Identifies as Woman<sup>a</sup> | 13.0 (4.8, 11.8-14.1) | 45.3 (7.0, 43.6-46.9) | **16.1 (3.5, 15.2-16.9)<sup>b</sup>** |
| Identifies as Man<sup>a</sup> | 11.3 (5.3, 9.0-13.6) | 48.1 (6.9, 45.1-51.2) | **17.8 (2.4, 16.8- 18.8)<sup>b</sup>** |
| Cohen's *d* Effect Size | 0.33 | 0.41 | 0.53 |
| Non-white identifying | 12.5 (5.4, 10.7-14.2) | 47.3 (7.2, 44.9-50.0) | 16.9 (3.4, 15.9-18.1) |
| White identifying | 12.6 (4.6, 11.4-13.9) | 44.9 (6.8, 43.0-46.8) | 16.2 (3.2, 15.2-17.0) |
| Cohen's *d* Effect Size | 0.02 | 0.34 | 0.25 |
| Never legally married | **12.2 (5.2, 11.1-13.4)<sup>b</sup>** | 46.1(7.1, 44.4-47.7) | 16.6 (3.3, 15.9-17.4) |
| Legally married, not separated | **14.3 (3.6, 12.7-16)<sup>b</sup>** | 44.9 (6.2, 41.9-47.8) | 16.0 (3.5, 14.3-17.7) |
| Cohen's *d* Effect Size | 0.42 | 0.17 | 0.18 |
| Close contact with person with MI | 12.8 (5.0, 11.7-13.8) | 45.2 (6.9, 43.6-46.7) | 16.1 (3.2, 15.3-16.7) |
| No close contact with person with MI | 11.3 (4.7, 8.4-14.2) | 51.2 (5.8, 47.4-54.9) | 19.4 (2.1, 18.0-20.8) |
| Cohen's *d* Effect Size | 0.29 | 0.88 | 1.07 |

<sup>a</sup>We had categories for learners identifying as non-binary as well as for self-description of gender identity however the cell-sizes were less than n=5 and are not reported here.
<sup>b</sup>Estimates in **bold** type font are statistically significant using independent samples t-tests (WHO-5 score at $P = 0.03$ (one-tailed), OMS-HC score at $P=0.02$ (two-tailed) and OMS-HC Subscale score at $P=0.01$ (two-tailed)).

Regarding the Self-Stigma of Mental Illness (SSMIS) scale, people who identified as non-white learners scored higher in the applying stigma of mental illness to oneself (Apply) subscale when compared to learners who identified as white (Geometric mean: 11.0 vs 8.8, *P* = 0.03).

In terms of substance use, 65% of participants reported using alcohol in the past 12 months while 38.1% reported the using marijuana in the last 12 months. Stimulants (4.0%) and LSD, mescaline, PCP, angel dust, mushrooms (4.0%) were less commonly used as were ecstasy, ketamine and MDMA (2.4%) and tranquilizers (1.6%).

### Qualitative (QUAL) – Interviews and Survey Comments

Table 3 shows the demographic characteristics of the interview participants.

**Table 3.** Demographic Characteristics of Interview Participants ^a^.

| Learner Type | Institution | Age | Gender identity | Marital Status | Living with Common Law | Ethno-racial Identity |
| --- | --- | --- | --- | --- | --- | --- |
| Medical student | Western Canada | 22 | Woman | Single | No | Chinese |
| Medical student | Western Canada | 25 | Man | Single | Yes | White |
| Medical student | Western Canada | 24 | Man | Single | No | White |
| Medical student | Eastern Canada | 24 | Woman | Single | No | South Asian |
| Medical student | Eastern Canada | 25 | Woman | Single | No | West Asian or Middle Eastern |
| Medical student | Eastern Canada | 23 | Woman | Single | No | Southeast Asian |
| Medical student | Eastern Canada | 26 | Woman | Single | Yes | White |
| Resident | Western Canada | 29 | Woman | Single | No | White |
| Resident | Western Canada | 29 | Woman | Married | No | White |

<sup>a</sup>The additional n=5 medical students and n=1 resident in this sample did not wish to disclose their demographic information.

#### Barriers and Enablers

The most prominent barriers to disclosure included fear and stigma. Fear was perceived as fear of judgment from peers, and fear of retribution or career consequences from those with more structural power such as preceptors or organizations themselves. Among medical student participants, many shared their fears of appearing as though they were substandard or not up to par as learners or future physicians. For example, one medical student participant described a fear “that I’m going to be a bad doctor somehow,” and also realized that this was due to their internalization of views of what would be considered a good doctor (M1). Among resident participants, however, there was more of a fear of negative consequences for their career rather than a fear of judgment. Several resident participants cited fear of future licensure applications including a survey comment that there is

> “Not just a fear of retribution for disclosing but knowing this happens from watching classmates disclose their struggles…watching residency programs force residents to take mental health leave for either real mental health concerns or as a power tool to intimidate residents into doing what the program wants or the residents permanently has a record of taking mental health leave. Taking leave from residency, or being diagnosed with a mental health condition, or being on/ having ever received treatment can affect future licensing and license applications and it can lead to mental health exclusions in future critical illness or disability insurance.” Survey Participant 01

Another resident participant stated,

> “And so I think I had a lot of – it’s not really fear of retribution, but just really recognizing that requesting any sort of accommodations in work would really negatively affect my learning experience in residency. And affect my job prospects down the road, because I think it would be very quickly construed as laziness or just maybe the inability to kind of unable to hack it, which is a very common trope.” R2

In terms of enablers of disclosure, the most prominent enabler was when learners had a preceptor that demonstrated genuine and empathic understanding, was non-judgmental, and preceptors who role modeled vulnerability. Supportive peers were also considered an enabler, along with having reassurance of anonymity and inclusive and transparent policies that would help learners know what would happen if they disclosed. Overall, participants noted that access to resources and an open and safe learning environment where disclosure is normalized serves as an enabler.

Enablers to self-disclosure were perceived differently in relation to participants’ views of themselves versus how they were perceived by others. Many participants shared that before they could disclose to others, they needed to give themselves permission to self-disclose. The concept of ‘permission to self’ included overcoming an individual’s internalized fears of being weak, less-than, or somehow not worthy of their place in the profession. Participants described working towards self-acceptance and perceiving a sense of agency or self-efficacy before feeling prepared to disclose to others. In contrast, when it came to enablers from one’s environment, an individual’s perceived comfort, perceived support, and perceived closeness to their peers were prominent enablers. Participants tended to disclose more to peers who were similar to them, however, some disclosed to preceptors and others to their organization’s wellness or wellbeing offices. There were also a few instances of accidental disclosure which occurred when participants sought accommodations and disclosed to acquire support that they perceived as necessary from their institution.

Most participants described a precipitant to their disclosure. The precipitant varied whether it was among peers, to their preceptor, or to their institution. Across all situations, there was a need for some type of facilitation or mediation before disclosure was elicited. In some circumstances it was a supportive preceptor who facilitated disclosure through gentle or non-judgmental questioning, in others it could be peers who made the discloser comfortable by demonstrating support, openness, and trust. Ultimately, motivations to disclose included a desire to support learning or gain accommodation, but also to help others or to role model vulnerability.

#### Consequences of Disclosure

Generally, participants described feeling positive after disclosure. They felt better and more confident about themselves. They also felt a sense of validation from their peers, closeness, and camaraderie that made them feel validated after their disclosure. Participants also noted that they felt that disclosing would help make them a better physician, have more empathy for patients, and be a better educator.

#### Variation in the Disclosure Journey

Overall, we found that there was tremendous variation by context, perceived identity, and diagnosis. Most individuals described unique and individual disclosure journeys and experiences. Among our participants, several described how their diagnosis or diagnoses had a unique influence on how they experienced disclosure. For example, one participant stated that they were not sensitive about their ADHD diagnosis but were concerned about “stigma around a diagnosis of bipolar” M5, while another disclosed their bipolar disorder but were hesitant to disclose their obsessive-compulsive disorder diagnosis M8. There was also a clear distinction between disclosure of mental illness versus substance use challenges. One participant stated that self-disclosure of depression and anxiety is “less awkward” than “something like substance abuse” M7, and a resident participant noted that,

> “If you’re struggling with substance abuse, and you’re taking morphine from the cabinets, then I think there’d be a greater risk disclose to the college, because they’ll certainly take your license away versus disclosing that to a peer may you know, they might try to work with you and allow you to keep going and just in a safer way.” R2

The degree that an individual’s self-perception of being different from others also varied. For example, one medical student stated that they felt like an “outsider” in the system and that their experience as an outsider influences how they feel about themselves and their discomfort with disclosure. (M7). Similarly, another participant shared those men have “their own particular challenges” with disclosure that are unique to others (M8), and one described how they feel “disclosures about my depression is kind of akin to disclosures about my sexuality…they are definitely interrelated.” M2.

#### Disclosure, Double Standards, and Internalization of Self-Stigmatizing Beliefs

A finding across all codes, groups, and participants related to how disclosure was understood and experienced uniquely in the context of medical training. Participants described a sense of hypocrisy, double standards, or duplicity. Several noted that application processes seemed to seek students who were well rounded and that narratives of adversity were often anticipated, yet, their experiences in medical training were discordant to their expectations and their conceptualization of what would make a good physician. One participant described cultural norms in medicine by stating,

> “But, this happens so much in medicine. We, like, talk about illnesses as if no one in the room could possibly have it. Like we talk about diabetes with this assumption, like, no one has diabetes. We talk about hepatitis as if no one has or has ever gotten hepatitis. We talk about OCD in this case and compulsions as if no one has compulsions in the room.” M8

Table 4 provides additional quotes from our qualitative findings.

**Table 4.** Representative Quotes from Qualitative Findings.

| **Barriers** | **Enablers** | **Consequences** | **Double Standards** |
| --- | --- | --- | --- |
| *Fear of judgment* | *Permission to self* | *Better about self* | *Internalized Stigma* |
| “Does this make me look like I’m weak or not good; that I’m going to be a bad doctor some how...I realized those are totally untrue, but I think that was just my internal monologue.” – M1 | “Disclosing is part of the whole process of acknowledging that happened to you and working to change your thinking.” – M9 | “Being able to disclose has taught me that I’ve accepted it; and the more I’m able to disclose that and share that the more accepting I am of myself, and you know...those struggles.” – M2 | “My own self stigma is just the self doubt part that is more of an internalized thing...an internalized fear...and just my own performance being affected.” – M6 |
| *Fear of retribution* | *Peers* | *Better doctor* | *Hypocrisy* |
| “It was just so embarrassing, so I didn’t want people to know if they didn’t have to know or if they weren’t going to help me. Just because I knew that would kind of change dynamics. And I didn’t want anything on a permanent record either.” - R1 | “It was the comfort of ‘I’m not in a room with strangers anymore, I’m in a room with people who know me, and I know them.’” – M6 | “I think it (disclosure) makes me able to ask more insightful questions and show some understanding that helps me build rapport with people quickly.” – M12 | “There’s learner wellness and wellbeing ...its almost a buzz word I find for the schools, but I don’t really see it put into action.” – M10 |
| *Culture of medicine* | *Preceptors* | *Better teacher* | *Culture of Medicine* |
| “When I think of students in medical school, it just feels like everyone is doing so well and everyone’s so smart and it seems like no one’s really struggling or having problems with anything.” – M4 | “She was just super warm...so open and curious...she asked a few questions, and it felt like it was going to be okay to say it.” – R3 | “I would love to teach in the future... and hope that I would be able to create an environment...that would make someone with similar difficulties...feel very comfortable and open to share that with me.” – R3 | “We are taught that empathy is the cornerstone of medicine...and then on the other hand we have the way that we treat each other, which is traditionally a very intimidating, hierarchical approach to teaching.” – R1 |

#### Overall Findings: The Journey from Fear to Confidence

Triangulation of quantitative and qualitative analysis suggests that disclosure was a complex process that appeared to become more challenging over time due to the internalization of negative attitudes about mental illness. OMS-HC scores showed resident physicians had more negative attitudes towards mental illness and disclosure. Overall findings also suggest that self-disclosure depended upon the degree of intersectional vulnerability of the learner’s identity and diagnosis. For example, the more intersections a learner’s identity encompassed, the more likely they perceived their self-disclosure of their mental illness or addition would be stigmatized. When looking at self-disclosure, people who identified as men had more negative attitudes than people who identified as women. Racialized learners scored higher on self-stigma. Qualitative data amongst participants with intersectional identities struggled in unique and varied ways not only according to other aspects of their identity such as gender and racialization, but also according to their perception of stigma related to their diagnosis. Quantitative estimates pertaining to substance use or addiction were unremarkable. Overall, findings suggested that disclosure journey was fraught with tensions, but ultimately lead to positive outcomes.

## Discussion

Our exploration of perceptions of self-disclosure among medical learners highlight the complexity of individual learners’ disclosure journey, and the internalization of pervasive norms regarding disclosure, vulnerability, and what constitutes a competent or effective physician. Disclosure experiences varied based on individuals’ intersectional identity and perceived diagnosis and appeared to become more difficult over time. However, disclosure was ultimately perceived as having positive outcomes for individuals and their beliefs about themselves and place in medicine.

Our findings suggests that despite being positive and complementary to participants’ professional identity formation, disclosure was difficult, fraught with tension, and varied according to their developmental stage in training. Our findings also support the relevance of employing an intersectional approach when assessing medical learner self-disclosure decision-making and wellbeing.^24^ We also learned that our participants scored within a range of poor overall wellbeing, and our findings complement other research which suggests that medical training is a unique risk factor for poor wellbeing which is further compounded by barriers to self-disclosure.^1,2,25,26^

Our participants confirmed the persistent double standards and perceived hypocrisy in medical education and that these double standards and fear of disclosure appears to worsen over time.^27^ For example, in our study resident OMS-HC scores and qualitative findings demonstrated fear of judgment among medical students become fear of retribution or adverse career consequences for learners. This is supported by the literature which shows any positive changes that may occur through best intended efforts of stigma reducing contact-based education with patients with mental illness early on in undergraduate training tend to decay over time as learners progress through the system.^28,29^ Yet, in a recent discourse analysis, participants also shared that discourses of vulnerability were in direct contrast to discourses of perfection, and that emotional suppression is taught both explicitly and implicitly to medical learners.^6^ Our findings therefore suggest that conventional approaches to support stigma reduction and foster disclosure may not be as effective in the context of medical education.

The system of medical education is in of itself a barrier to self-disclosure, yet self-disclosure is perceived as positive once it has happened based on learners’ perspectives. Our study suggests that the existing approaches that might foster disclosure in conventional settings will be insufficient in a medical education context unless longstanding norms and perfectionism are addressed. Structural changes and accountability through regulatory and accreditation standards may be necessary. For example, accrediting bodies may want to consider how medical schools address stigma and normalize self-disclosure by preceptors and learners alike which can be an active ingredient in learner self-disclosure. ^3,17,29^

The process of facilitating disclosure was varied but ultimately shaped by all actors and influences, highlighting the importance of everyone’s role in medical learners’ sphere of influence addressing stigma. Disclosure was varied, yet it happened in all contexts and with individuals. Some participants disclosed to peers, others to preceptors, and some to their institution itself. The key enabler included perceptions regarding support and safety. Furthermore, the importance of social support can not be understated given that participants who were married reported higher wellbeing scores as well as qualitative findings pertaining to support. This form of social support was highlighted by peers, preceptors, and family members alike.

Lastly, given the intersectional aspects of how medical learners perceive their identity and how such factors influence disclosure, we recommend that medical schools and residency programs consider the importance of disaggregated socio-demographic data about learner’s intersectionality when planning and implementing wellness initiatives and interventions. For example, people who identify as men may have more negative attitudes towards self-disclosure due to societal norms and racialized learners may be more likely to experience self-stigma based on their own cultural norms about mental illness and potential fear of double discrimination.^30,31^ which ultimately has downstream impacts for health equity. Our findings should help inform any interventions to reduce stigma by ensuring that other aspects of social identity are addressed, named, and space is created for thoughtful conversations about double discrimination.

Our study had several limitations. First, we collected data solely on the disclosure of mental illness or addiction rather than a holistic perspective of learner wellbeing^26^ taking into consideration other aspects of health such as financial stability, physical illness etc. which can have an impact on mental health or addiction as well as overall wellbeing. Second, measuring stigma related to mental illness or addiction may always have the risk of social desirability bias which may skew the data. Given that the data we found on several of our measures was positively skewed, we are unable to ascertain whether participant attitudes were positive or whether participants wanted to appear socially desirable due to fear of retribution. Third, our sample size for the quantitative arm of the study was small and the quantitative results should be interpreted with caution. We recognize the limitations of recruiting individuals for a study with a sensitive and challenging topic such as self-disclosure. Having said this, there is enough evidence given the triangulation of the quantitative and qualitative results of a signal in the noise suggesting further research into intersectional identities in medical learners and the role they play in self-disclosure of mental illness or addiction. Last, we used a long battery of questionnaires which may have imposed response burden for participants and may have contributed to a smaller sample size of survey participants and missing data.

Future research will need to address the impact of identity markers on medical learner experiences of training and the impact of the learning environment on self-disclosure and wellbeing, for example how the disclosure occurs i.e., peer-to-peer vs. to individuals of power vs. unintended disclosures for accommodation.). Further exploration is also warranted on how best to tackle the structural and cultural barriers in medical education to foster self-disclosure in an ethical manner as well as the role of mentorship in facilitating disclosure.

## Conclusions

The individual process of disclosure is complex and appeared to become more challenging over time due to the internalization of negative attitudes about mental illness. Intersectional vulnerability in medical learners warrants further consideration because it may modulate disclosure. Fear of disclosure is an important factor shaped by the learning environment that has implications for learner wellbeing, patient care and health equity.

*Acknowledgments:* The authors wish to thank all the participants for their time and efforts. *Funding/Support:* This study was funded by the Association of Medical Education of Europe (AMEE) Medical Education Research Grant.

*Other disclosures:* The authors have nothing to disclose.

*Ethical approval:* This research was approved by the Conjoint Health Research Ethics Board at the University of Calgary (ID:20-1149) and the Office of Human Research Ethics (OHRE) at Western University (ID: 116634).

*Previous presentations:*

Kassam A, Antepim B, & Sukhera J. (2023). Exploring perceptions of self-disclosure in medical education. *International Congress on Academic Medicine*, Quebec City, Canada, April 13-18, 2023.

Kassam A, Antepim B, & Sukhera J. (2022). Exploring Perceptions of Self-Disclosure in Medical Education. *Transform MedEd Conference*, London, England, November 11-12, 2022.

## Data Availability

All data produced in the present study are available upon reasonable request to the authors.

