## Supplemental Material for "Disclosure and Double Standards: A Mixed Methods Study of Self-Disclosure of Mental Illness or Addiction Among Medical Learners"

| **Scale** | **Abbreviation** | **Construct Measured** | **Number of Items** | **Scoring** | **Subscale Information** |
| --- | --- | --- | --- | --- | --- |
| Opening Minds Scale for Healthcare Providers | OMS-HC | Stigma towards Mental Illness | 20 | A 5-point Likert scale was used, and response options were 1 = Strongly disagree, 2 = Disagree, 3 = Neither agree nor disagree, 4 = Agree and 5 = Strongly agree. Scores could range from 20 to 100 and a lower score indicated less stigma. Items 3, 8, 9, 10, 11, 15, 19 required reverse scoring. Subscale scores are based on the 12-item OMS-HC. The scoring of the attitudes of healthcare providers towards people with mental illness subscale (7 items) may range from 7 (least stigmatizing) to 35 (most stigmatizing) while the scoring for the attitudes towards disclosure of a mental illness subscale (5 items) may range from 5 (least stigmatizing) to 25 (most stigmatizing). | Two subscales: 1) Attitudes Towards Mental Illness and 2) Attitudes Towards Disclosure of Mental Illness |
| World Health Organization (WHO) Wellbeing Scale | WHO-5 | General wellbeing and depression | 5 | The raw score is calculated by totaling the five answers. The raw score ranges from 0 to 25, 0 representing worst possible and 25 representing best possible quality of life. To obtain a percentage score ranging from 0 to 100, the raw score is multiplied by 4. A percentage score of 0 represents worst possible, whereas a score of 100 represents best possible quality of life. | Not applicable. |
| Self-Stigma of Mental Illness Scale | SSMIS | Self stigma associated with mental illness | 40 | Responses use a nine-point agreement scale (9=strongly agree). Scores were determined by summing only the five items for each subscale that remained in the shortform, yielding a range of scores between 5 and 45 for each of the four subscales. A score below 13 indicates poor wellbeing and is an indication for testing for depression under ICD-10. | Four subscales: 1) Awareness of mental illness and public attitudes, 2) Agreement with attitudes, 3) Application of mental illness to oneself, and 4) Having a mental illness and harm to one’s self esteem. |
